## Supplementary figures S1-S13 and Supplementary Tables T1-T3 for "Longitudinal development of sex differences in the limbic system is associated with age, puberty and mental health"

### Supplementary File

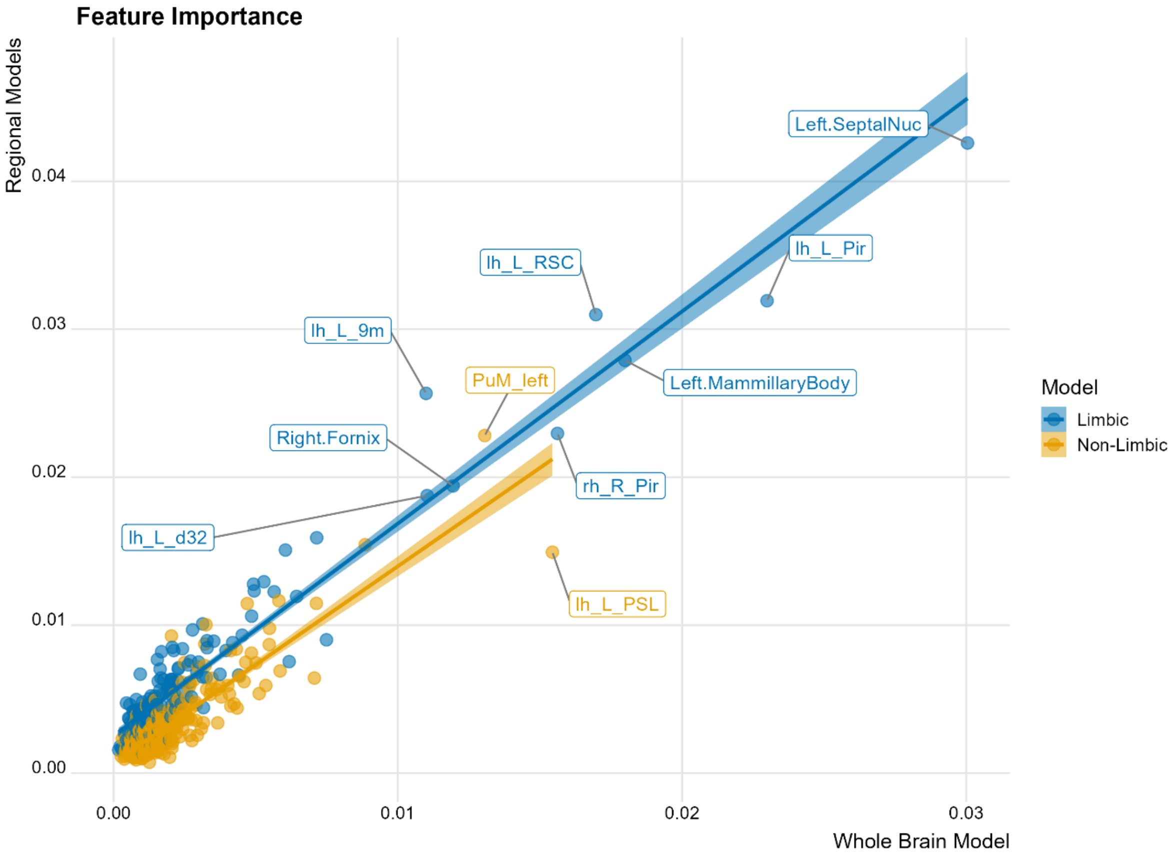


**Figure S1. Comparisons of feature contributions between regionally constrained and whole brain models.** Features contributing the most to the limbic and non-limbic models also contributed the most to the whole brain model. Regions contributing the most to the limbic model were the septal nuclei, the piriform cortex, subregions of the anterior and posterior cingulate cortex and the mammillary body. Regions contributing the most to the non-limbic model were the peri-Sylvian language area and the left medial pulvinar.

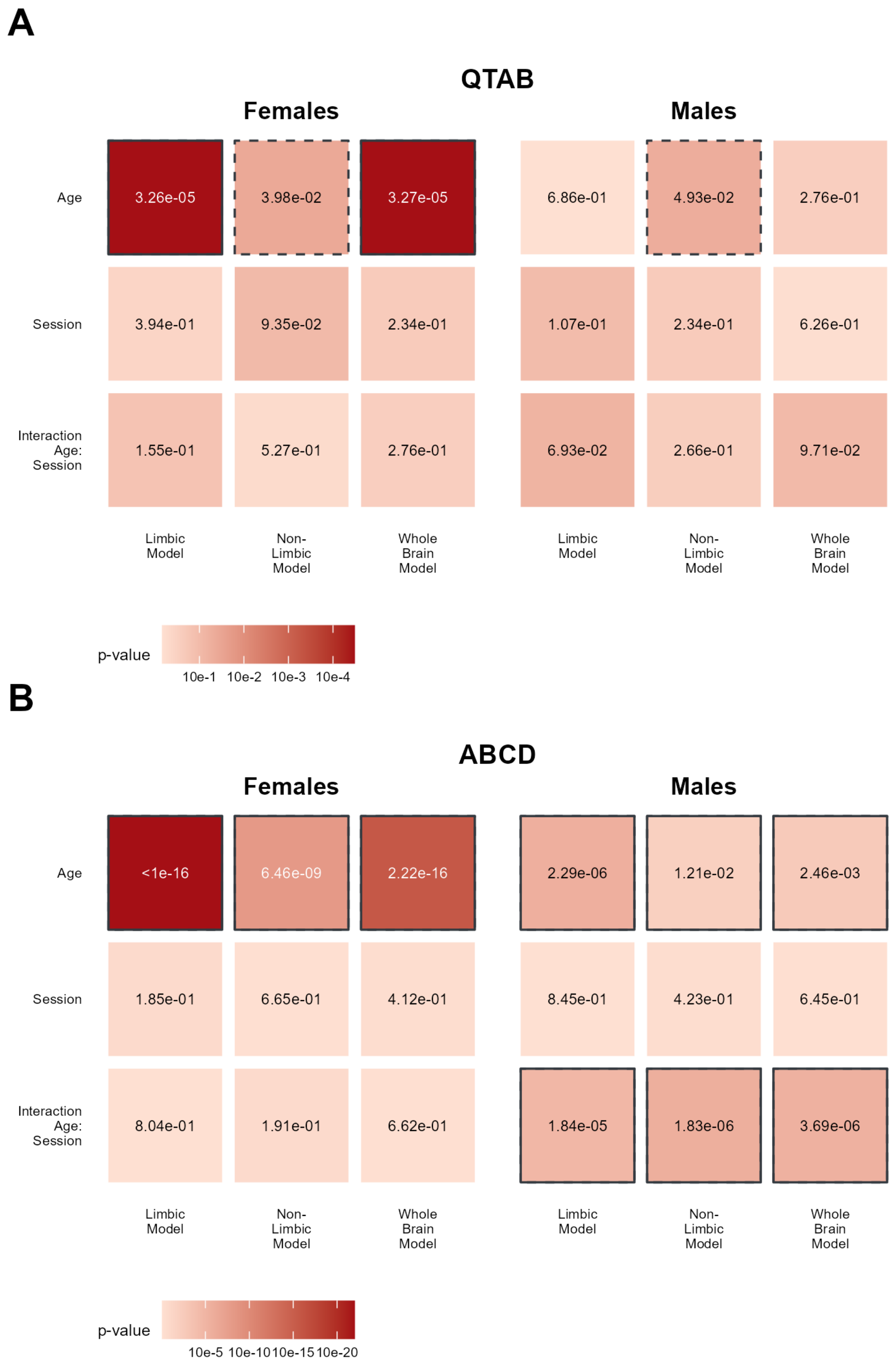
A
QTAB
Females
Males
Age
Session
Interaction Age: Session
3.26e-05
3.98e-02
3.27e-05
6.86e-01
4.93e-02
2.76e-01
3.94e-01
9.35e-02
2.34e-01
1.07e-01
2.34e-01
6.26e-01
1.55e-01
5.27e-01
2.76e-01
6.93e-02
2.66e-01
9.71e-02
Limbic Model
Non-Limbic Model
Whole Brain Model
p-value
10e-1 10e-2 10e-3 10e-4
B
ABCD
Females
Males
Age
Session
Interaction Age: Session
<1e-16
6.46e-09
2.22e-16
2.29e-06
1.21e-02
2.46e-03
1.85e-01
6.65e-01
4.12e-01
8.45e-01
4.23e-01
6.45e-01
8.04e-01
1.91e-01
6.62e-01
1.84e-05
1.83e-06
3.69e-06
Limbic Model
Non-Limbic Model
Whole Brain Model
p-value
10e-5 10e-10 10e-15 10e-20

**Figure S2. Class probabilities are significantly associated with age, with stronger association in females compared to males.** The maps depict the p-values associated with the F-statistics presented in Figure 1 of the main manuscript for each dataset, derived with Linear Mixed Effect (LME) models. Solid borders denote significance after correction for multiple comparisons ($p < .017$), dashed border denote nominally significant values that do not survive correction for multiple comparisons ($p < .05$)

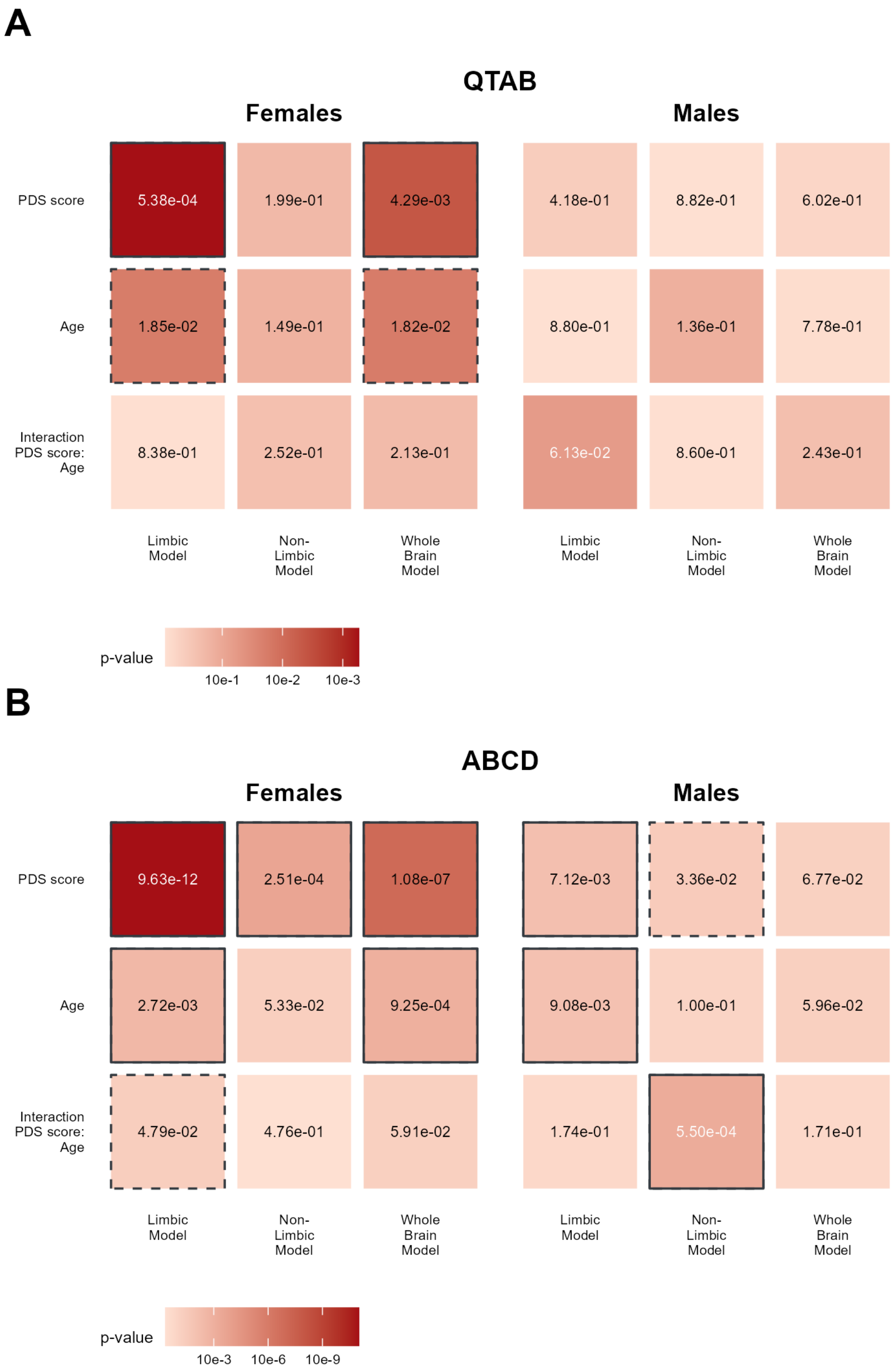
A
QTAB
Females
Males
PDS score
5.38e-04
1.99e-01
4.29e-03
4.18e-01
8.82e-01
6.02e-01
Age
1.85e-02
1.49e-01
1.82e-02
8.80e-01
1.36e-01
7.78e-01
Interaction PDS score: Age
8.38e-01
2.52e-01
2.13e-01
6.13e-02
8.60e-01
2.43e-01
Limbic Model
Non-Limbic Model
Whole Brain Model
Limbic Model
Non-Limbic Model
Whole Brain Model
p-value
10e-1
10e-2
10e-3
B
ABCD
Females
Males
PDS score
9.63e-12
2.51e-04
1.08e-07
7.12e-03
3.36e-02
6.77e-02
Age
2.72e-03
5.33e-02
9.25e-04
9.08e-03
1.00e-01
5.96e-02
Interaction PDS score: Age
4.79e-02
4.76e-01
5.91e-02
1.74e-01
5.50e-04
1.71e-01
Limbic Model
Non-Limbic Model
Whole Brain Model
Limbic Model
Non-Limbic Model
Whole Brain Model
p-value
10e-3
10e-6
10e-9

**Figure S3. Class probabilities are associated with average PDS score with a pattern of strongest results in females and in the limbic system across datasets.** The maps depict the p-values associated with the F-statistics presented in Figure 2 of the main manuscript for each dataset, derived with Linear Mixed Effect (LME) models. Solid borders denote significance after correction for multiple comparisons ($p < .017$), dashed borders denote nominally significant values that do not survive correction for multiple comparisons ($p < .05$).

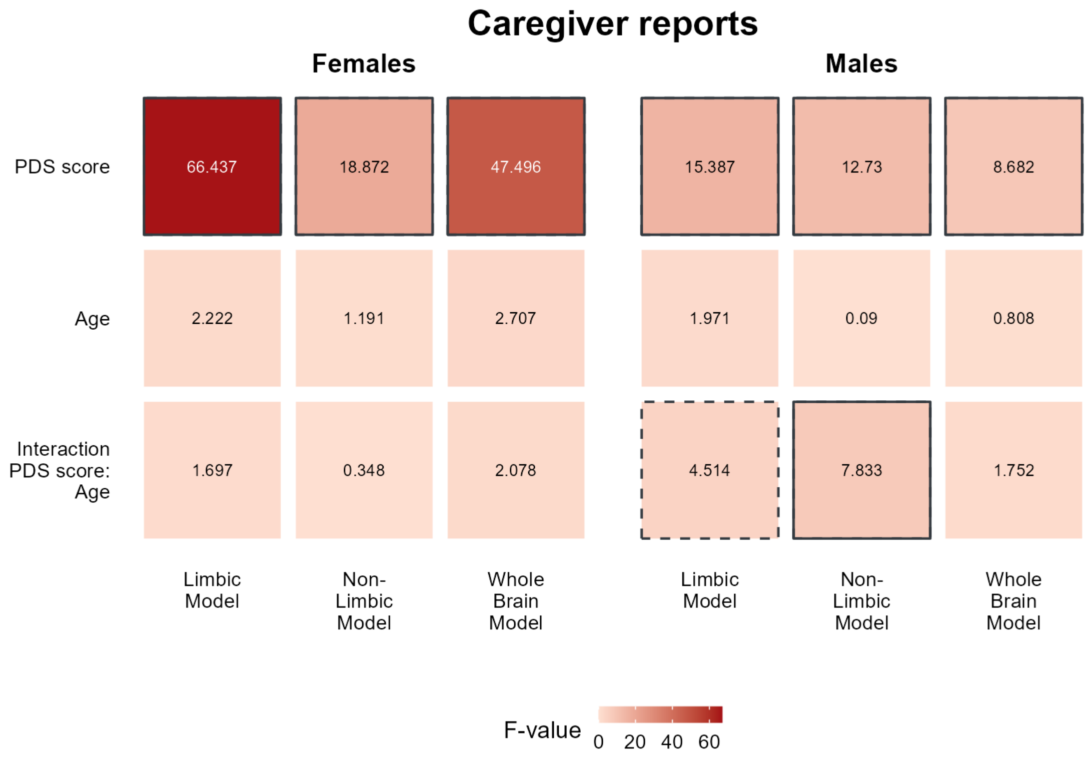


**Figure S4. Caregiver's reports Pubertal Developmental Scale (PDS) score is associated with class probabilities in ABCD.** In line with the findings obtained with self-reported PDS, limbic class probabilities showed strongest associations with PDS score with a pattern of stronger associations in females compared to males. Solid borders denote significance after correction for multiple comparisons ($p < .017$), dashed borders denote nominally significant values that do not survive correction for multiple comparisons ($p < .05$).

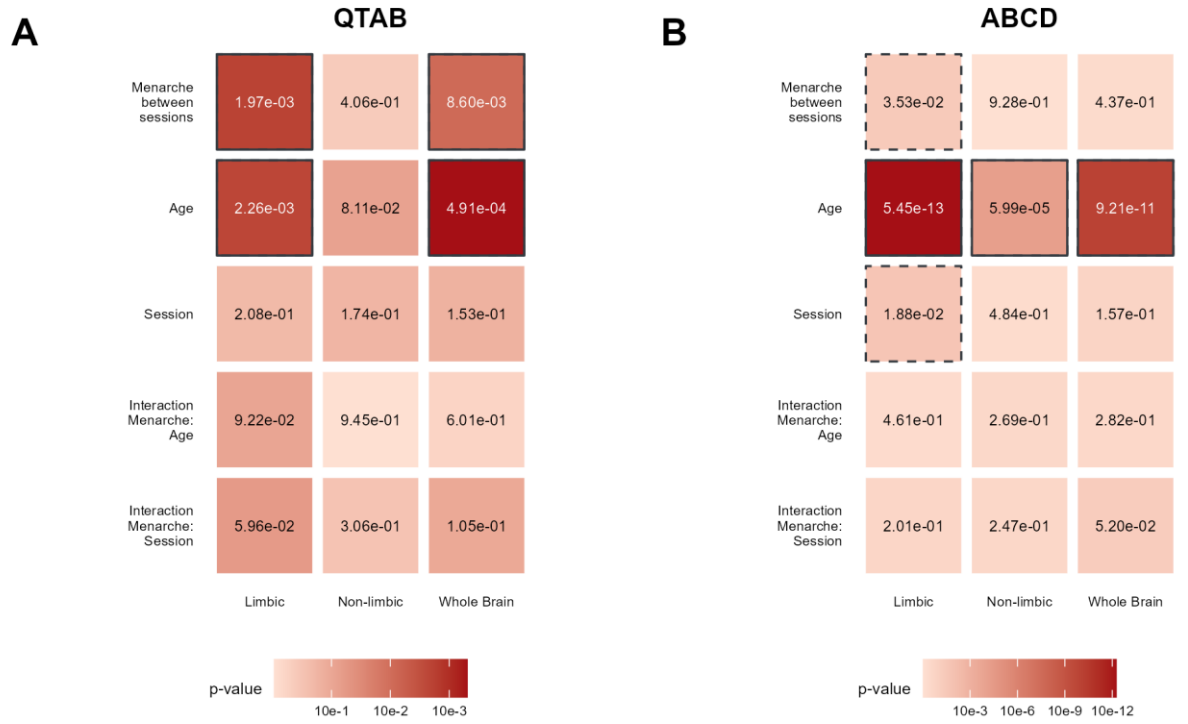


**Figure S5. Limbic class probabilities are sensitive to menarche onset.** The maps depict the p-values associated with the F-statistics presented in Figure 3 of the main manuscript for each dataset, derived with Linear Mixed Effect (LME) models. Solid borders denote significance after correction for multiple comparisons ($p < .017$), dashed borders denote nominally significant values that do not survive correction for multiple comparisons ($p < .05$).

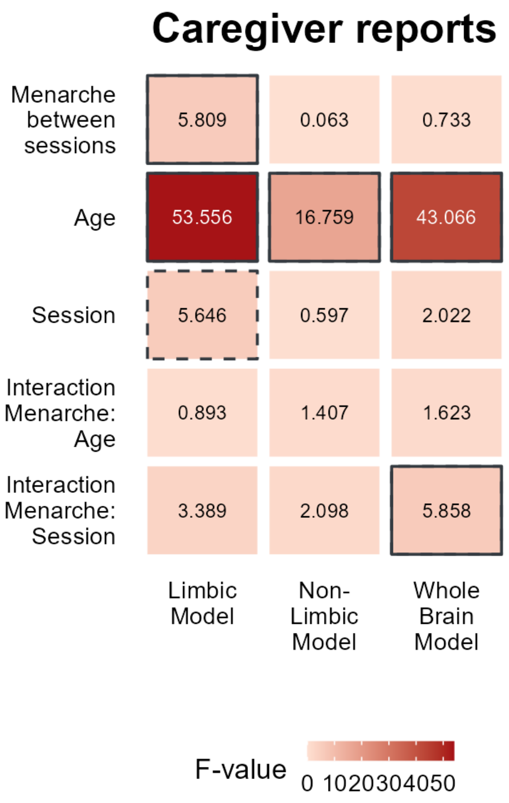


**Figure S6. Menarche between sessions derived from caregiver's reports is associated with limbic class probabilities in ABCD.** Solid borders denote significance after correction for multiple comparisons ($p < .017$), dashed borders denote nominally significant values that do not survive correction for multiple comparisons ($p < .05$).

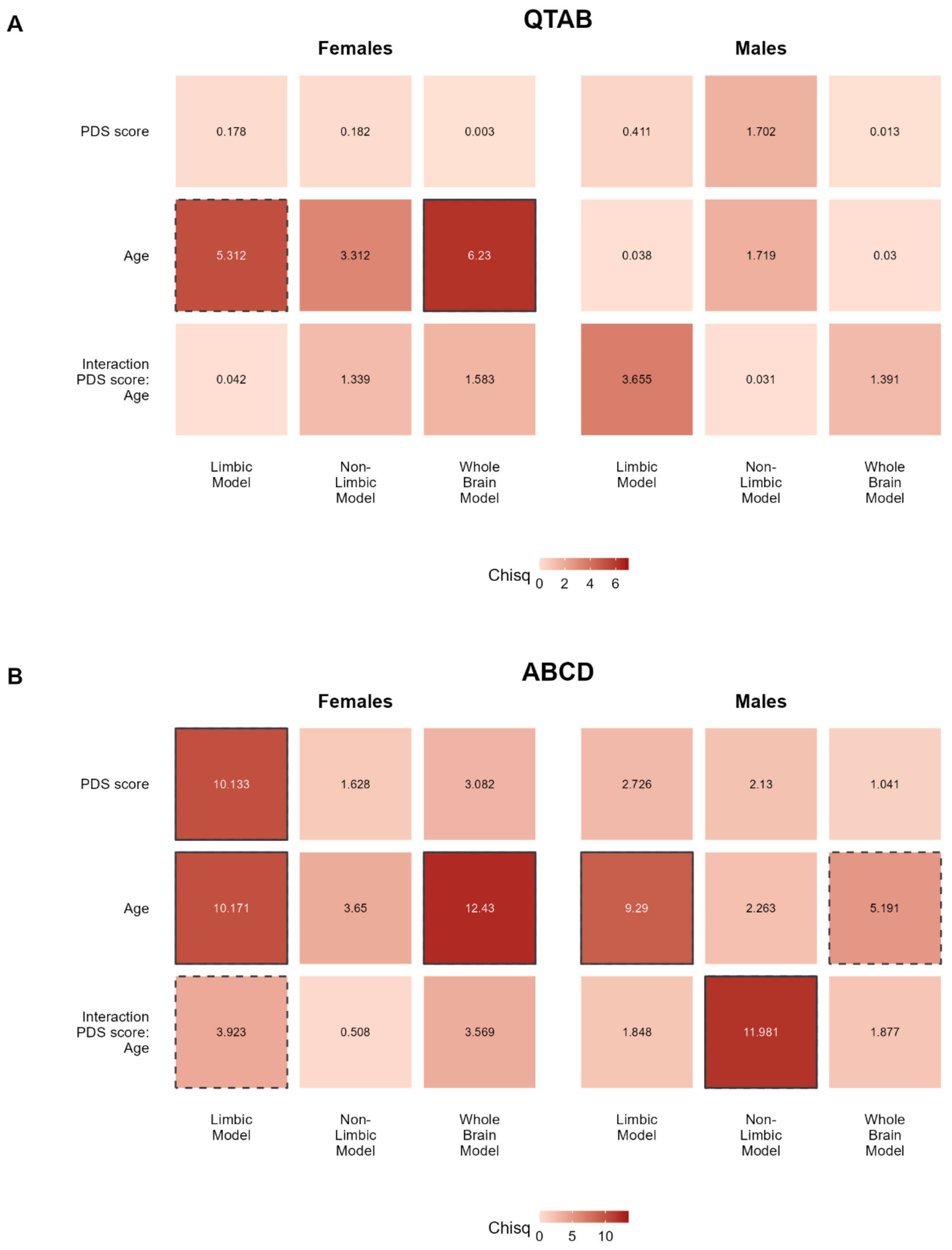


**Figure S7. Relative contributions of age and PDS score to the associations with class probabilities, by ANOVA type-II.** A) In QTAB, age explained most of the variance in the associations with class probabilities for both sexes. B) In ABCD, limbic class probabilities in females are significantly associated with PDS beyond age, while age has strongest effects for the other models and in males. Solid borders denote significance after correction for multiple comparisons ($p < .017$), dashed borders denote nominally significant values that do not survive correction for multiple comparisons ($p < .05$).

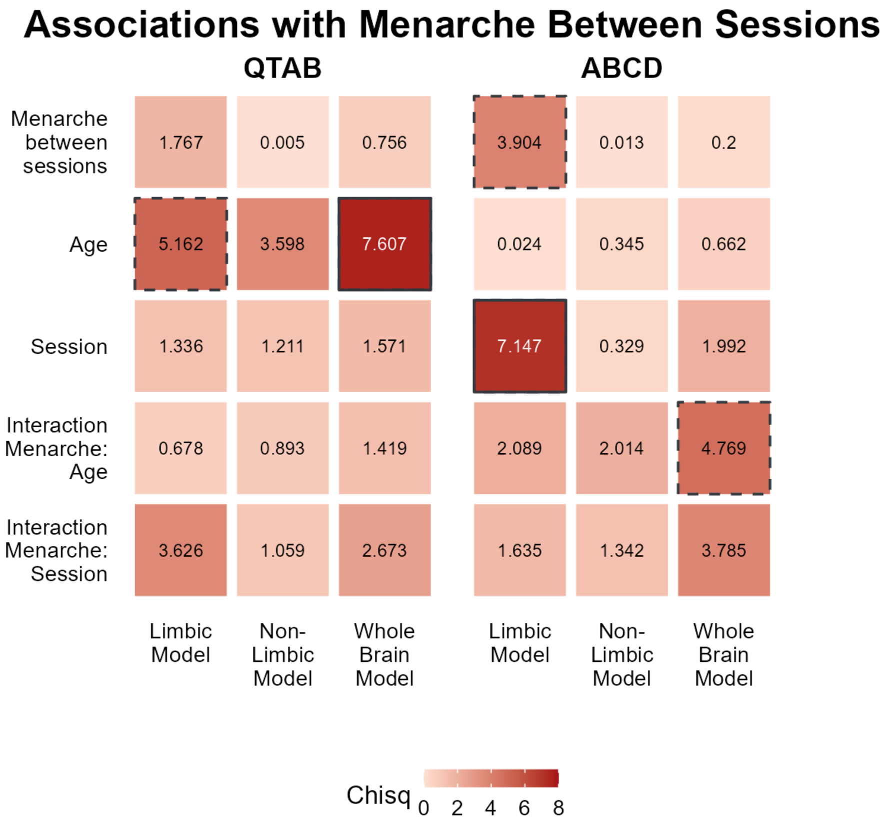


**Figure S8. Relative contributions of age and menarche onset between sessions with ANOVA type-II.** A) In QTAB, age explained most of the variance, while B) a significant association of menarche onset between sessions was found in ABCD. Solid borders denote significance after correction for multiple comparisons ($p < .017$), dashed borders denote nominally significant values that do not survive correction for multiple comparisons ($p < .05$).

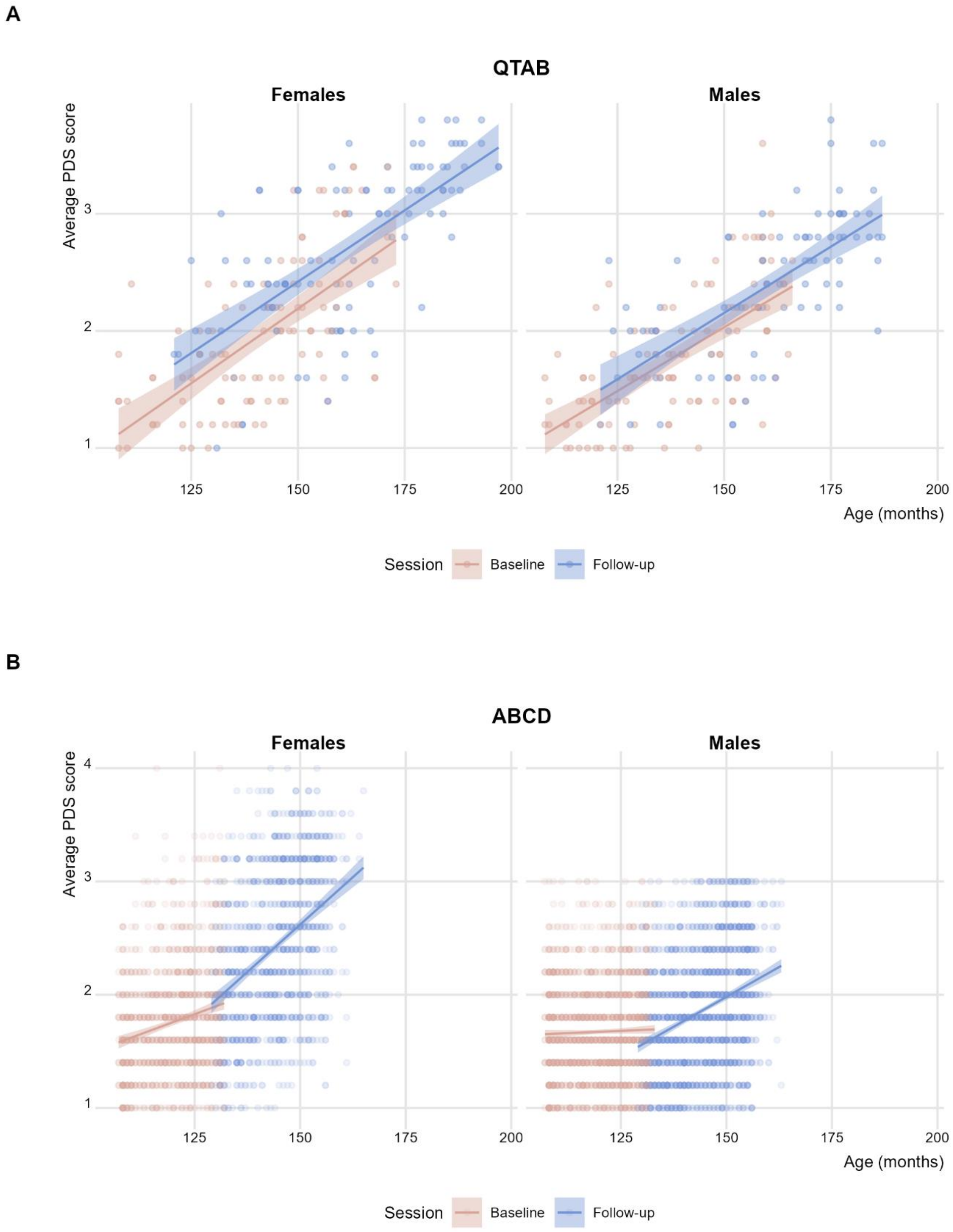


**Figure S9. Correlation between PDS score and age.** A) In QTAB, strong significant correlations were found at each session in both females and males. No significant difference in correlations between sessions were found. B) In ABCD, females showed a significant correlation between age and PDS score for both sessions, while males showed a significant correlation only at follow-up. Moreover, a significant difference between correlation at baseline and follow-up was found in each sex.

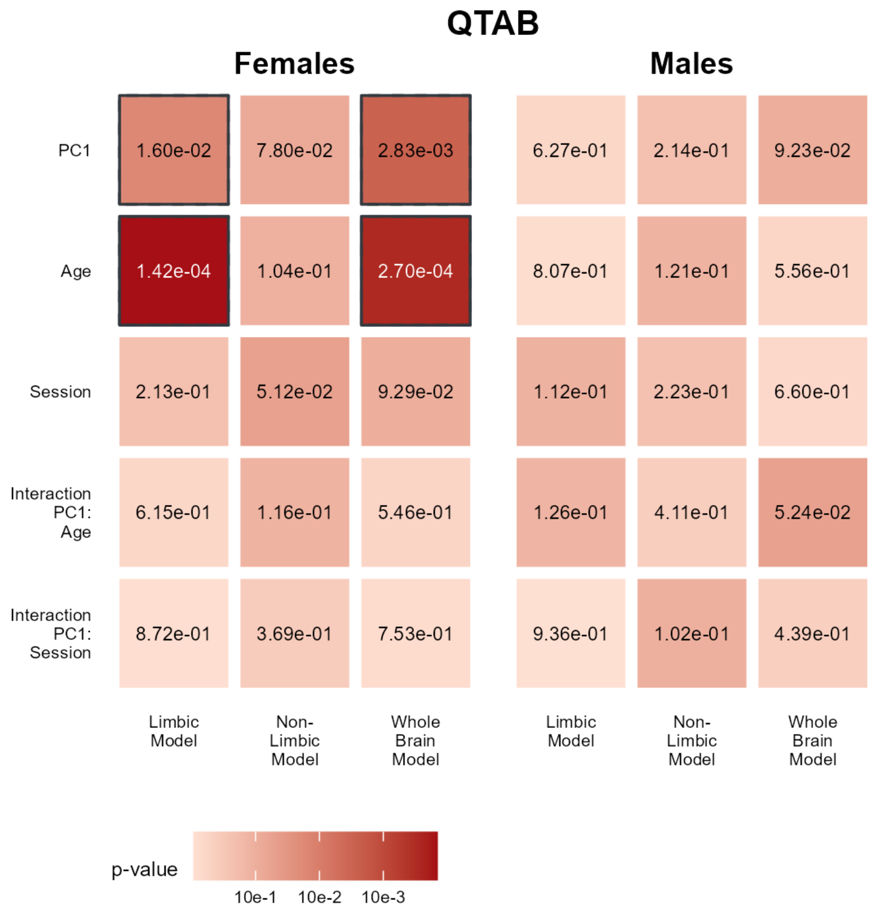


**Figure S10. Class probabilities are associated with mental health in a sex-specific manner.** The maps depict the p-values associated with the F-statistics presented in Figure 4 of the main manuscript for each dataset, derived from Linear Mixed Effect (LME) models. Solid borders denote significance after correction for multiple comparisons ($p < .017$).

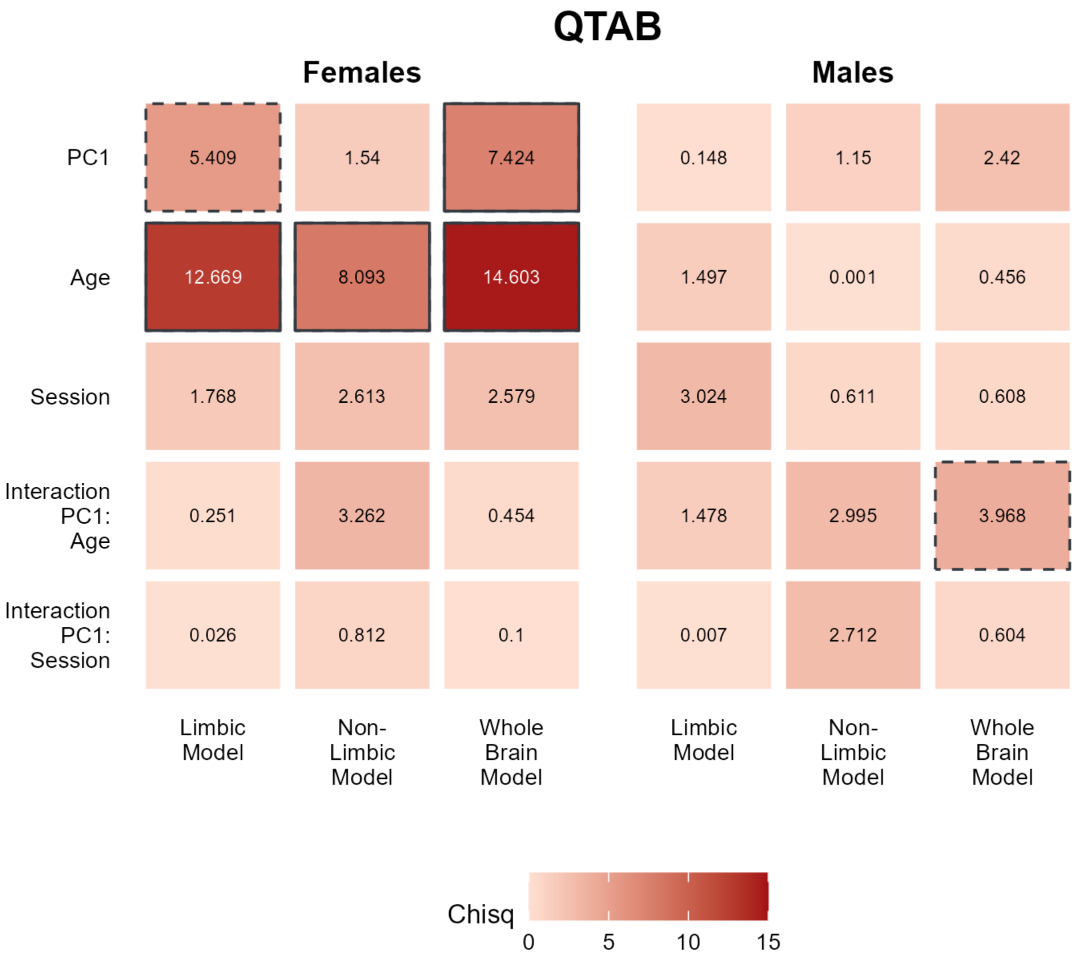


**Figure S11. Class probabilities are associated with both mental health status beyond age in a sex-specific manner.** Anova type-II results showed significant association of whole brain and limbic class probabilities with mental health score beyond age in females, while no significant effect was found in males. After correction for multiple comparisons only the whole brain effect survived. Solid borders denote significance after correction for multiple comparisons ($p < .017$), dashed borders denote nominally significant values that do not survive correction for multiple comparisons ($p < .05$).

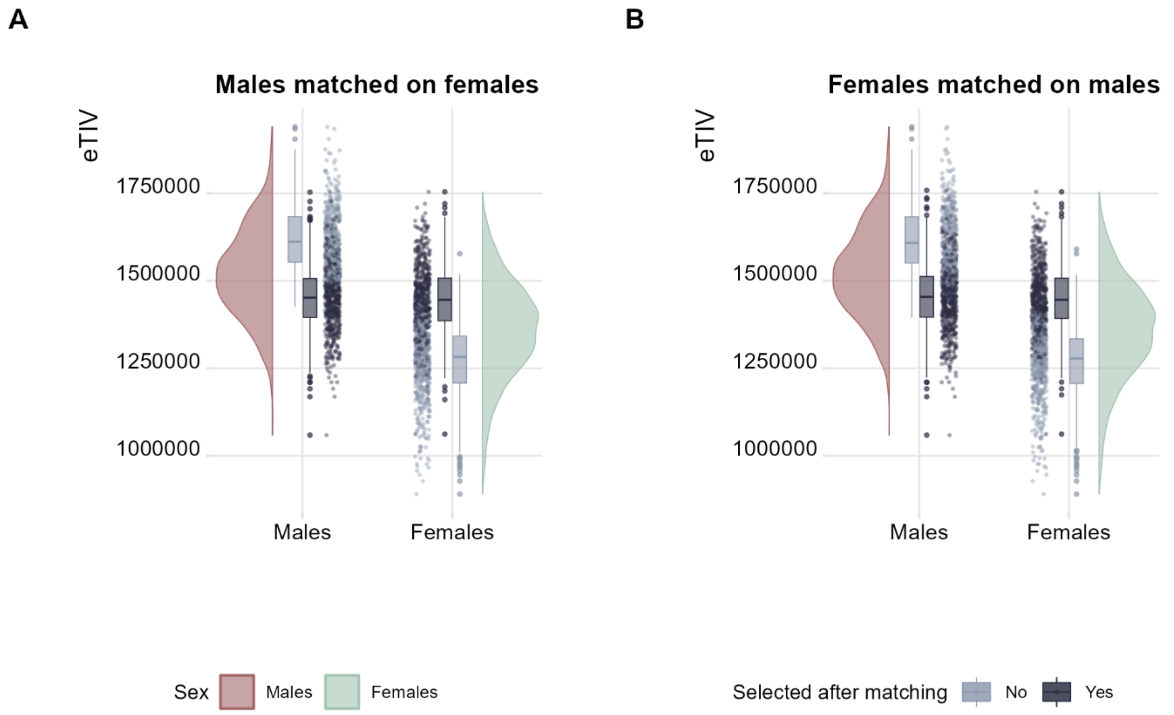


**Figure S12. Matching procedure using each of the two sexes as reference leads to comparable Estimated Total Intracranial Volume (eTIV) distribution.** A) eTIV distribution when females are taken as reference, corresponding to the training set used in the main paper. B) Supplementary analysis using males as reference leads to similar distribution.

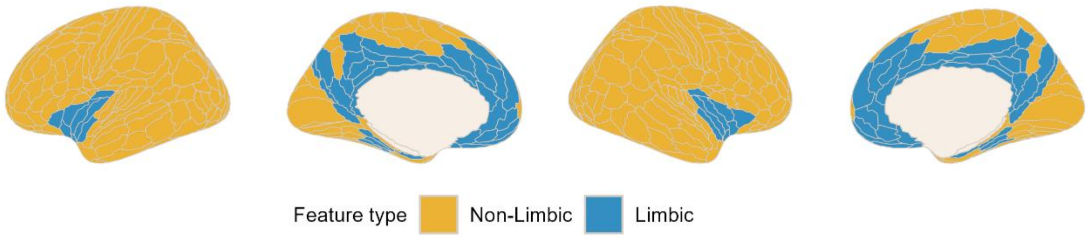


**Figure S13. Cortical distribution of limbic and non-limbic features.**

### Supplementary Tables

| | Total Sample | | |
|---|---|---|---|
| | Tot | Females | Males |
| N | 2054 | 1086 | 968 |
| age | 179 ± 44.5 | 181 ± 45.0 | 178 ± 43.9 |
| eTIV | 1436981 ± 154645.9 | 1361084 ± 136872.0 | 1522131 ± 126595.0 |
| | Selected | | |
| | Tot | Females | Males |
| N | 1132 | 566 | 566 |
| age | 174 ± 44.6 | 174 ± 44.5 | 174 ± 44.7 |
| eTIV | 1449042 ± 98151.0 | 1447216 ± 99663.0 | 1450869 ± 96670.0 |
| | Excluded | | |
| | Tot | Females | Males |
| N | 922 | 520 | 402 |
| age | 187 ± 43.4 | 188 ± 44.3 | 184 ± 42.0 |
| eTIV | 1422173 ± 202685.0 | 1267332 ± 107026.0 | 1622465 ± 90666.0 |

**Table T1. Training sample characteristics before and after the matching procedure for the selected and excluded participants.** The eTIV distributions support that the matching procedure successfully removed the eTIV difference between males and females, while still staying within the bounds of a reasonably representative sample. Mean ± standard deviation (SD) of age (expressed in months) and eTIV.

| | | | | Baseline | | Follow-up | |
|---|---|---|---|---|---|---|---|
| | | | | Females | Males | Females | Males |
| QTAB | | | | | | | |
| | Age | | | 137 ± 16.87 | 136 ± 15.79 | 158 ± 19.14 | 155 ± 17.54 |
| | N | | | | | | |
| | | Tot | | 192 | 200 | 152 | 138 |
| | | PDS | | 185 | 179 | 145 | 119 |
| | | Menarche | | 110 | - | 110 | - |
| | | | No menarche | 64 | - | 64 | - |
| | | | Between sessions | 46 | - | 46 | - |
| | | Mental Health | | 192 | 200 | 152 | 138 |
| ABCD | | | | | | | |
| | Age | | | 119 ± 7.41 | 119 ± 7.49 | 143 ± 7.78 | 144 ± 7.75 |
| | N | | | | | | |
| | | Tot | | 3629 | 4121 | 3478 | 4070 |
| | | PDS | | 1173 | 1846 | 1159 | 1841 |
| | | Menarche | | 1091 | - | 1091 | - |
| | | | No menarche | 677 | - | 677 | - |
| | | | Between sessions | 414 | - | 414 | - |

**Table T2. Included participants for each association analysis in the two longitudinal samples.** Age is expressed in months and reported as mean ± standard deviation (SD)

**Table T3. List of limbic structures with the correspondent feature in Freesurfer**

| Structure | FreeSurfer Feature | Segmentation |
|---|---|---|
| Anterior Cingulate Cortex | 33pr | Glasser atlas |
| | p24pr | |
| | a24pr | |
| | p24 | |
| | a24 | |
| | p32pr | |
| | a32pr | |
| | d32 | |
| | p32 | |
| | s32 | |
| | 8BM | |
| | 9m | |
| | 10v | |
| | 10r | |
| | 25 | |
| Orbitofrontal Cortex | OFC | |
| | pOFC | |
| Insula | MI | |
| | AVI | |
| | AAIC | |
| | Ig | |
| | PI | |
| | PoI1 | |

| Structure | FreeSurfer Feature | Segmentation |
|---|---|---|
| | PoI2 | |
| Piriform Cortex | Pir | |
| Entorhinal Cortex | EC | |
| Parahippocampal Area | PHA1 | |
| | PHA2 | |
| | PHA3 | |
| Posterior Cingulate Cortex | DVT | |
| | ProS | |
| | POS1 | |
| | POS2 | |
| | RSC | |
| | v23ab | |
| | d23ab | |
| | 31pv | |
| | 31pd | |
| | 31a | |
| | 23d | |
| | 23c | |
| | PCV | |
| Hippocampus | Hippocampal_tail | Hippocampal Subfields |
| | subiculum-body | |
| | CA1-body | |
| | subiculum-head | |
| | hippocampal-fissure | |
| | presubiculum-head | |
| | CA1-head | |
| | presubiculum-body | |
| | parasubiculum | |

| Structure | FreeSurfer Feature | Segmentation |
|---|---|---|
| | molecular_layer_HP-head | |
| | molecular_layer_HP-body | |
| | GC-ML-DG-head | |
| | CA3-body | |
| | GC-ML-DG-body | |
| | CA4-head | |
| | CA4-body | |
| | fimbria | |
| | CA3-head | |
| | HATA | |
| Amygdala | Lateral-nucleus | Nuclei of Amygdala |
| | Basal-nucleus | |
| | Accessory-Basal-nucleus | |
| | Anterior-amygdaloid-area-AAA | |
| | Central-nucleus | |
| | Medial-nucleus | |
| | Cortical-nucleus | |
| | Corticoamygdaloid-transitio | |
| | Paralaminar-nucleus | |
| Anterior and Dorsomedial Thalamic Nuclei | AV | Thalamic Nuclei |
| | LD | |
| | MDl | |
| | MDm | |
| Nucleus Accumbens | Nucleus-Accumbens | ScLimbic |
| Hypothalamus | HypoThal-noMB | |
| Fornix | Fornix | |
| Mammillary Body | MammillaryBody | |
| Forebrain | Basal-Forebrain | |

| Structure | FreeSurfer Feature | Segmentation |
|---|---|---|
| Septal Nuclei | SeptalNuc | |
